# Individual factors influencing public’s perceptions about the importance of COVID-19 immunity certificates: a cross-sectional online questionnaire survey in the UK

**DOI:** 10.1101/2021.11.12.21266256

**Authors:** Corina-Elena Niculaescu, Isabel Karen Sassoon, Irma Cecilia Landa-Avila, Ozlem Colak, Gyuchan Thomas Jun, Panagiotis Balatsoukas

**Author notes:** Correspondence to Dr. Panagiotis Balatsoukas.

## Abstract

**Objectives:** To assess what were the main individual factors influencing people’s perception of the importance of using COVID-19 immunity certificates.

**Design:** Cross-sectional online survey.

**Setting:** Nationally representative survey in the UK, conducted on the 3^rd^ of August 2021.

**Participants:** Responses from 534 participants, aged 18 and older, residents of the UK.

**Interventions:** This was a cross-sectional survey and each participant replied to the same set of questions.

**Primary outcome measure and independent variables:** The primary outcome measure (dependent variable) was the participants’ perceived importance of using immunity certificates, computed as an index of six items. The following individual drivers were used as the independent variables: a) personal beliefs about COVID-19 (using constructs adapted from the Health Belief Model), b) personal views on vaccination, c) willingness to share immunity status with service providers, and d) variables related to respondents’ lifestyle and socio-demographic characteristics.

**Results:** Perceived importance of immunity certificates was higher among respondents who felt that contracting COVID-19 would have a severe negative impact on their health *(β=0.2564, p=0.0000)* and felt safer if vaccinated *(β =0.1552, p=0.0000)*. The prospect of future economic recovery positively influenced perceived importance of immunity certificates. Respondents who were employed or self-employed *(β=-0.2412, p=0.0010)*, or experienced an increase in income after the COVID-19 pandemic *(β=-0.1287, p=0.0020)* perceived less important the use of immunity certificates compared to those who were unemployed or had retired or those who had experienced reduction in their income during the pandemic.

**Conclusions:** The findings of our survey suggest that more vulnerable members in our society (unemployed or retired and those believing that COVID-19 would have a severe impact on their health) and people who experienced a reduction in income during the pandemic perceived the severity of not using immunity certificates in their daily life as higher.

## 1. Introduction

While quite a few studies have tried to explore the role of different individual characteristics on attitudes towards vaccination[1–4], there is little known about their role on people’s attitudes towards immunity certificates. Immunity certificates, and their terminological variation like immunity passports or vaccine passports, have been at the centre of controversy as their value polarises opinions amongst academics, policy makers and the general public. Both perceived benefits of and concerns about immunity certificates have been reported in the literature. For example, preserving freedom of movement[5], re-opening the economy and reducing the risk of infection[6,7] are some frequently reported benefits, while loss of autonomy[8–13], legal challenges[14,15], risk of fraud[10] and digital exclusion[6,16,17] represent some of the most prominent concerns. This knowledge is useful in order to understand drivers and hinders of implementing immunity certificates in general. However, empirical evidence is needed to understand how different individual factors and characteristics may influence the prevalence of those drivers or hinders. Production of this knowledge is important to help us understand how perceptions around immunity certificates are influenced by individual characteristics and use this insight to inform policy making and implementation strategies for services around immunity certification, e.g. by helping identify those who are more in need of using immunity certificates[18,19].

The aim of the present paper is to report the findings of a UK wide online questionnaire survey assessing the role of different individual factors on perceived importance of using immunity certificates. Specifically, we examined the following types of individual factors personal beliefs about COVID-19, views on vaccination, willingness to share their immunity status, lifestyle, and socio-demographic characteristics. Throughout this paper we use the term “immunity certificate” to describe a service that allows individuals with antibodies of SARS-COV-2, obtained through past infection or after a full course of vaccination, to evidence their immunity status.

## 2. Methods

### 2.1. Sample Design

Our analysis is based on a cross-sectional dataset obtained from an online anonymous questionnaire survey, designed using the online platform OnlineSurveys *(onlinesurveys.ac.uk)*. Responses were collected using Prolific *(prolific.co.uk)* on the 3^rd^ of August 2021. Respondents were demographically representative of the UK population in terms of gender, age, and ethnicity. We excluded 20 participants who failed the attention checks, and one duplicate responder, resulting in a final sample of 534 respondents. All participants were 18 or older and were compensated for their participation in the study with £1.75/person. All materials including dataset, statistical codes, questionnaire survey and ethics approval can be accessed on OSF *(https://osf.io/jubv6/.)*

### 2.2. Main variables measure

#### 2.2.1. Perceived importance of using immunity certificates (Primary Outcome)

Perceived importance using immunity certificates was the computed index of six items each measuring a different area where the use of immunity certificates could impact people’s lives. A screenshot of the six items used is presented in Figure 1. Table 1 presents summary statistics for all variables used. These six items were informed by the findings of a series of focus groups and interviews investigating public’s concerns about the risks and unintended consequences of immunity certificates[18]. Responses to these items were measured on a 5-point Likert scale from (1 – “Strongly disagree” to 5 – “Strongly agree).

**Figure 1.**
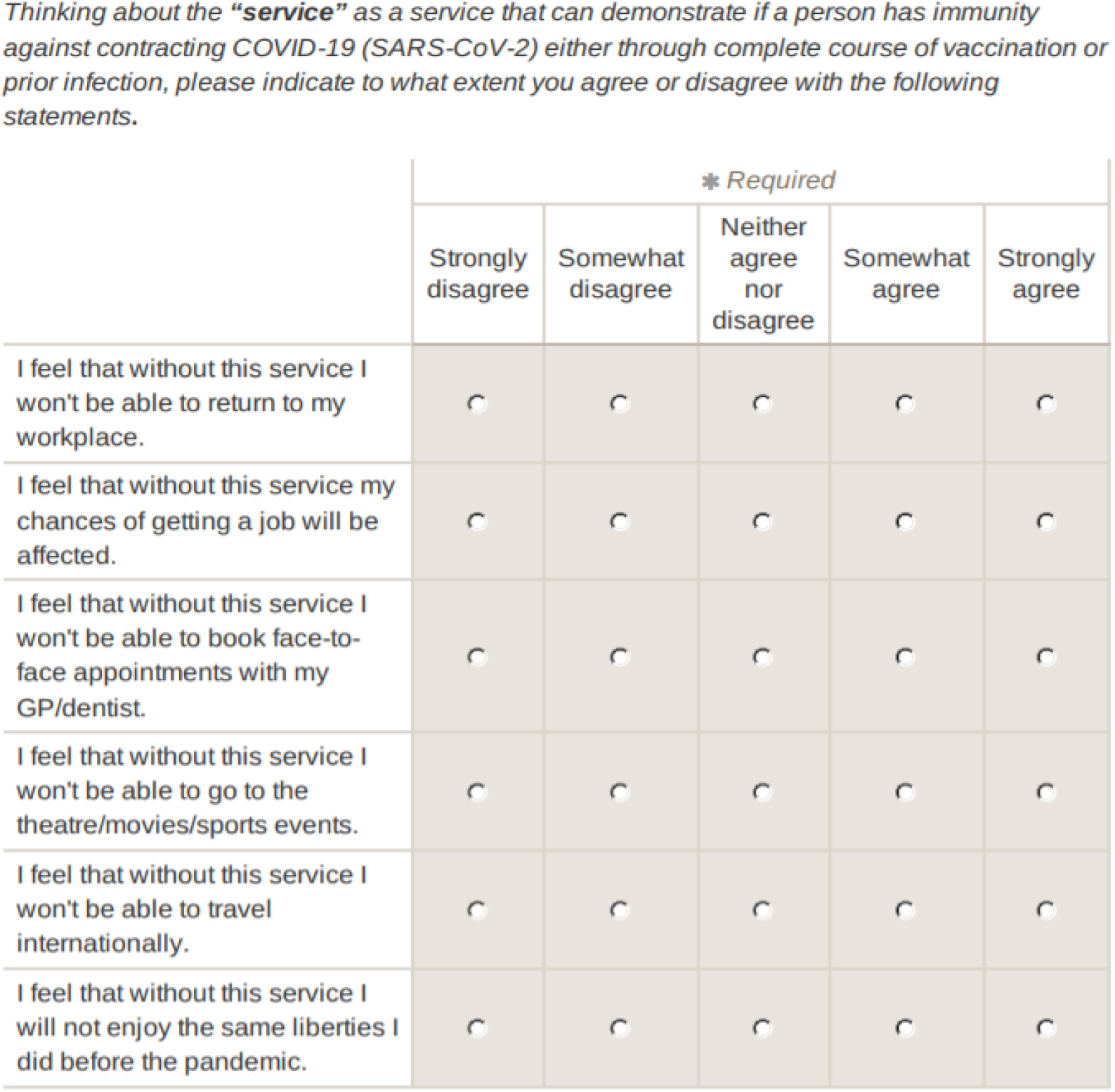
Perceived Severity of not Using Immunity Certificates Survey Questions *(https://osf.io/jubv6/.)*

**Table 1.** Summary statistics of HBM measures, vaccine views, lifestyle variables, willingness to share immunity status with service providers

| HBM Measures | Items | Mean | Median | Std. Dev | Min | Max | Alpha |
| --- | --- | --- | --- | --- | --- | --- | --- |
| Perceived Importance of Using Immunity Certificates | I feel that without this service I won't be able to return to my workplace. | 2.4476 | 2.0000 | 1.1558 | 1 | 5 | 0.8485 |
|  | I feel that without this service my chances of getting a job will be affected. | 2.5918 | 3.0000 | 1.1631 | 1 | 5 |  |
|  | I feel that without this service I won't be able to book face-to-face appointments with my GP/dentist. | 2.8371 | 3.0000 | 1.2455 | 1 | 5 |  |
|  | I feel that without this service I won't be able to go to the theatre/movies/sports events. | 3.2715 | 4.0000 | 1.1636 | 1 | 5 |  |
|  | I feel that without this service I won't be able to travel internationally. | 3.912 | 4.0000 | 1.1252 | 1 | 5 |  |
|  | I feel that without this service I will not enjoy the same liberties I did before the pandemic. | 3.6667 | 4.0000 | 1.1692 | 1 | 5 |  |
| Perceived COVID-19 Susceptibility | I am at risk of getting COVID-19 (SARS-CoV-2). | 3.5243 | 4.0000 | 1.1255 | 1 | 5 | 0.7095 |
|  | It is likely that I will get COVID-19 (SARS-COV-2). | 2.9401 | 3.0000 | 1.0122 | 1 | 5 |  |
|  | Individuals in my household are at risk for getting COVID-19 (SARS-COV-2). | 3.4438 | 4.0000 | 1.131 | 1 | 5 |  |
|  | I feel knowledgeable about my risk of getting COVID-19 (SARS-COV-2). | 4.1255 | 4.0000 | 0.746 | 1 | 5 |  |
| Perceived COVID-19 Severity | I believe that COVID-19 (SARS-COV-2) is a severe health problem in general. | 4.2266 | 4.0000 | 0.9662 | 1 | 5 | 0.7061 |
|  | If I get COVID-19 (SARS-COV-2) I will get sick. | 3.7247 | 4.0000 | 0.9749 | 1 | 5 |  |
|  | If I get COVID-19 (SARS-COV-2) I will die. | 2.1386 | 2.0000 | 0.9227 | 1 | 5 |  |
|  | If I get COVID-19 (SARS-COV-2) other members in my household will get sick. | 3.5824 | 4.0000 | 1.012 | 1 | 5 |  |
| Perceived Benefits of Immunity Certificates | This service will make me feel safe only if immunity is obtained through complete course of vaccination. | 3.2809 | 3.0000 | 1.141 | 1 | 5 | 0.6045 |
|  | This service will make me feel safe only if immunity is obtained through past COVID-19 (SARS-CoV-2) infection. | 2.4326 | 2.0000 | 0.9734 | 1 | 5 |  |
|  | This service will facilitate economic recovery. | 3.5506 | 4.0000 | 1.054 | 1 | 5 |  |
|  | This service will facilitate social gatherings in closed spaces without restrictions (e.g. wearing masks, limits on number of people who can gather). | 3.7154 | 4.0000 | 0.9922 | 1 | 5 |  |
| Perceived Barriers of using Immunity Certificates | I'm afraid that my data will be passed on to third parties without my consent or commercialized. | 3.0281 | 3.0000 | 1.2883 | 1 | 5 | 0.3691 |
|  | This service will be difficult for me to use if available only on smartphones / tablets. | 1.9307 | 2.0000 | 1.1913 | 1 | 5 |  |
|  | This service will be difficult for me to access if offered exclusively in English. | 1.2809 | 1.0000 | 0.6982 | 1 | 5 |  |
| Hopelessness after COVID-19 | Mental wellbeing after COVID-19 | 2.6685 | 3.0000 | 0.7606 | 1 | 5 |  |
|  | Net Income after COVID-19 | 2.8221 | 3.0000 | 0.8445 | 1 | 5 |  |
| Vaccine Views | I am not convinced that the vaccine will protect me against COVID-19 (SARS-CoV-2). | 2.3034 | 2.0000 | 1.2212 | 1 | 5 |  |
|  | I feel worried about people who have received a non-UK approved vaccine entering the country. | 2.6292 | 3.0000 | 1.2055 | 1 | 5 |  |
| Lifestyle | Travel internationally for business | 1.3820 | 1.0000 | 0.7428 | 1 | 4 |  |
|  | Travel internationally for leisure | 2.6330 | 3.0000 | 0.9230 | 1 | 4 |  |
|  | Travel internationally to visit family and/or friends | 2.0243 | 2.0000 | 1.0293 | 1 | 4 |  |
|  | Book accommodation (hotels, Airbnb etc.) | 2.8333 | 3.0000 | 0.8879 | 1 | 4 |  |
|  | Attend sports events | 2.0300 | 2.0000 | 0.9583 | 1 | 4 |  |
|  | Go to the theatre or movies | 2.8015 | 3.0000 | 0.8521 | 1 | 4 |  |
|  | Visit museums, galleries and other cultural exhibitions or festivals | 2.7828 | 3.0000 | 0.8090 | 1 | 4 |  |
|  | Go to a pub, restaurant, club or coffee shop for a meal or drink. | 3.4045 | 4.0000 | 0.7634 | 1 | 4 |  |
|  | Care for or visit someone who lives in a care home. | 1.5112 | 1.0000 | 0.8850 | 1 | 4 |  |
| Willingness to share immunity | Theatre/cinema/gallery | 3.2921 | 4.0000 | 1.3998 | 1 | 5 |  |
| status with service providers | Pub/restaurant. | 3.2228 | 4.0000 | 1.4159 | 1 | 5 |  |
|  | GP/dentist | 4.4700 | 5.0000 | 0.9219 | 1 | 5 |  |
|  | Hospitality sector | 3.4663 | 4.0000 | 1.3717 | 1 | 5 |  |
|  | Sports event | 3.3015 | 4.0000 | 1.4012 | 1 | 5 |  |
|  | Airport/airline | 3.8764 | 4.0000 | 1.2538 | 1 | 5 |  |
| Hopelessness after COVID-19 | Mental wellbeing after COVID-19 | 2.6685 | 3.0000 | 0.7606 | 1 | 5 |  |
|  | Net Income after COVID-19 | 2.8221 | 3.0000 | 0.8445 | 1 | 5 |  |

The distribution of responses for each item is presented in Figure 2. Subsequently, we observed that the internal reliability of the six items was high (0.8485)[20] (Table 1). Therefore, we measured the overall perceived importance of using immunity certificates, by creating the index *Certificate Severity*. This index was computed as the average score amongst its six component items, and it is a continuous variable taking values between 1 and 5 [21].

**Figure 2.**
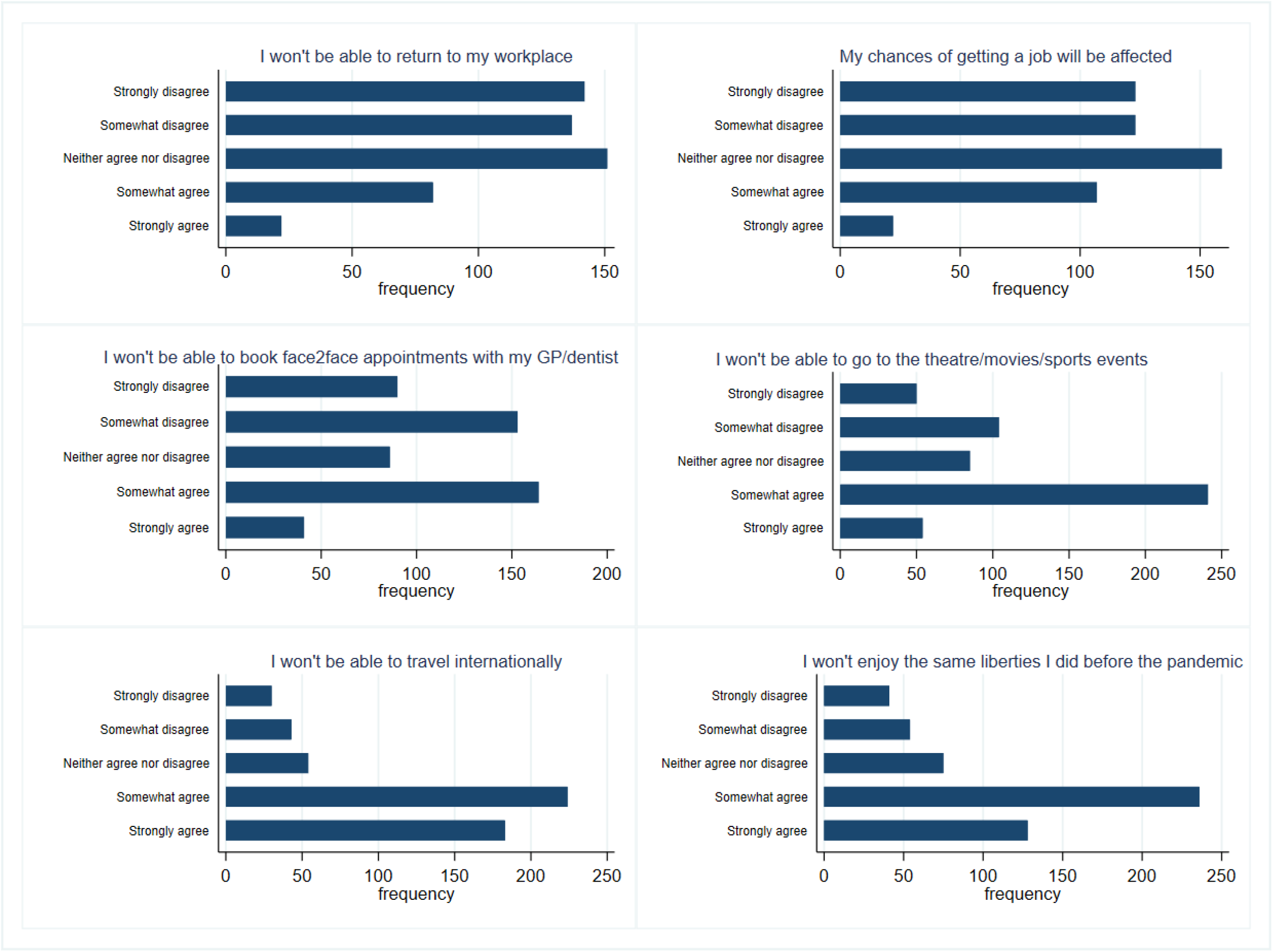
Distribution of responses across perceived severity of not using immunity certificates *(https://osf.io/jubv6/.)*

### 2.3. Independent variables

#### 2.3.1. Personal beliefs about COVID-19

We measured respondents’ personal beliefs about COVID-19 using four constructs adapted from the Health Belief Model (HBM)[22] and tailored to the needs of the present study. The detailed description of the items, summary statistics and internal reliability measures are presented in Table 1. Each item was rated on a 5-point Likert scale from 1 (“Strongly disagree”) to 5 (“Strongly agree”). First, we measured perceived COVID-19 susceptibility using three items adapted from [2] and one item from [23]. Second, we measured perceived COVID-19 severity through four items adapted from [2]. Perceived COVID-19 susceptibility measures respondents perceived risk of contracting the SARS-Cov-2 virus, while perceived COVID-19 severity represents the perceived severity of negative health consequences if the respondent were to contract the virus. Third, we measured perceived barriers from using immunity certificates with three items referring to data safety and accessibility (smartphone availability and language). Finally, we measured perceived benefits of using immunity certificates through four items covering safety, economic recovery, and return to social gatherings.

As presented in Table 1, perceived COVID-19 susceptibility and perceived COVID-19 severity display a Cronbach’s Alpha of 0.7 or higher, suggesting good internal consistency. Therefore, we created an index (*Perceived COVID-19 Susceptibility, Perceived COVID-19 Severity*) for each of these constructs by averaging the items within the constructs[24,25]. For perceived barriers and perceived benefits of using immunity certificates we used the individual items in our analysis, without transforming these into indices, as their Cronbach alpha was lower than 0.7[20].

#### 2.3.2. Vaccination Views

At the time when our study was conducted approximately 75% of the UK’s adult population had been vaccinated[26]. Therefore, instead of employing the traditional HBM constructs of measuring intention to get vaccinated, vaccination barriers or perceived severity of COVID-19 vaccines, we asked three questions on vaccination views that our previous qualitative research showed were common concerns among both fully vaccinated and not vaccinated individuals[18]. As such we constructed three questions about respondents’ perceived vaccine effectiveness, worries about non-UK approved vaccines, and feeling of safety around vaccinated people. Each item was rated on a 5-point Likert scale from 1 (“Strongly disagree”) to 5 (“Strongly agree”).

#### 2.3.3. Prior to COVID-19 Lifestyle

We asked a series of lifestyle-related questions to determine if respondents’ habits before the COVID-19 outbreak had an effect, if any, on the primary outcome measure. Lifestyle questions measured the frequency with which respondents engaged with a series of social activities using a 4-point Likert scale ranging from 1 (“Never”) to 4 (“Very often”). The complete list of questions is presented in Table 1. In summary these measured the frequency with which respondents travelled internationally, booked accommodation when travelling, attended sports events, went to theatres/movies or visited other cultural events, went to pubs, restaurants and other dinning venues, or visited a health care setting (for example, visited someone in a care home). Like in the case of the questions about vaccination views, the lifestyle questions were informed by the findings of our qualitative research conducted between February and July 2021[18].

#### 2.3.4. Willingness to share Immunity Status with Different Service Providers

Respondents were asked to rate their level of agreement in sharing their immunity status with different service providers on a 5-point Likert scale ranging from 1 (“Strongly disagree”) to 5 (“Strongly agree”). The types of service providers for which respondents had to rate their level of agreement included their GP/dentist, airport/airline, hospitality sector (e.g. hotels and other booked accommodation), theatre/cinema/gallery, sports event, pub/restaurant/nightclub.

#### 2.3.5. Socio-demographics

Summary statistics for the socio-demographic variables used in this study are presented in Table 2. In addition to the representative gender, age and ethnicity variables we also recorded data about respondents’ geographic location in the UK (urban or rural), accommodation arrangements (e.g. living alone or in shared accommodation), employment status, education, and whether or not the respondent had a disability.

**Table 2.** Demographic characteristics of sample

|  |  | **Freq.** | **Percent** | **Cum.** |
| --- | --- | --- | --- | --- |
| **Gender** | Female | 277 | 51.87% | 51.87% |
|  | Male | 254 | 47.57% | 99.44% |
|  | Self-define | 1 | 0.19% | 100% |
|  | Prefer not to say | 2 | 0.37% | 99.81% |
| **Age** | 18 – 23 | 77 | 14.42% | 14.42% |
|  | 24 – 29 | 51 | 9.55% | 23.97% |
|  | 30 – 39 | 95 | 17.79% | 41.76% |
|  | 40 – 49 | 87 | 16.29% | 58.05% |
|  | 50 – 59 | 95 | 17.79% | 75.84% |
|  | 60 – 69 | 109 | 20.41% | 96.25% |
|  | 70 or older | 20 | 3.75% | 100% |
| **Ethnicity** | Asian | 34 | 6.37% | 6.37% |
|  | Black | 20 | 3.75% | 10.11% |
|  | Hispanic/Latino | 3 | 0.56% | 10.67% |
|  | Mixed | 15 | 2.81% | 13.48% |
|  | Other | 8 | 1.50% | 14.98% |
|  | South Asian | 12 | 2.25% | 17.23% |
|  | White | 442 | 82.77% | 100% |
| **Region** | East Midlands | 42 | 7.87% | 7.87% |
|  | East of England | 35 | 6.55% | 14.42% |
|  | London | 81 | 15.17% | 29.59% |
|  | Northeast | 32 | 5.99% | 35.58% |
|  | Northern Ireland | 11 | 2.06% | 37.64% |
|  | Northwest England | 58 | 10.86% | 48.50% |
|  | Scotland | 37 | 6.93% | 55.43% |
|  | South-East England | 87 | 16.29% | 71.72% |
|  | Southwest of England | 43 | 8.05% | 79.78% |
|  | Wales | 19 | 3.56% | 83.33% |
|  | West Midlands | 45 | 8.43% | 91.76% |
|  | Yorkshire and the Humber | 44 | 8.24% | 100% |
| **Area** | Rural | 166 | 31.09% | 31.09% |
|  | Urban | 368 | 68.91% | 100% |
| **Accommodation** | Living alone | 87 | 16.29% | 16.29% |
|  | Living in shared accommodation | 54 | 10.11% | 26.40% |
|  | Living with other family members | 382 | 71.54% | 97.94% |
|  | Other | 11 | 2.06% | 100% |
| **Employment** | Employed/Self-employed | 340 | 63.67% | 63.67% |
|  | Retired | 97 | 18.16% | 81.84% |
|  | Unemployed | 97 | 18.16% | 100% |
| **Education** | A level (or equivalent) | 130 | 24.34% | 24.34% |
|  | GCSE (or equivalent) | 80 | 14.98% | 39.33% |
|  | Postgraduate degree | 95 | 17.79% | 57.12% |
|  | Undergraduate degree | 175 | 32.77% | 89.89% |
|  | Vocational | 54 | 10.11% | 100% |
| **Disability** | No | 467 | 87.45% | 87.45% |
|  | Prefer not to say | 6 | 1.12% | 88.58% |
|  | Yes | 61 | 11.42% | 100% |
| **All** |  | 534 | 100% |  |

The COVID-19 pandemic and subsequent lockdown measures have been difficult for many people leading to deceased mental wellbeing[27–32], unemployment and/or lower income[33,34]. Therefore, to control for the possibility of attitudes towards the primary outcome measure streaming from feelings of hopelessness, we measured perceived mental wellbeing and net income now compared to before the beginning of the pandemic using a 5-point Likert scale ranging from 1(“Much worse”/”Much lower”) to 5(“Much better”/”Much higher”).

### 2.4. Statistical Analysis

In order to address our research questions, we employed a multiple stepwise linear regression analysis using *Certificate Severity* (i.e. respondents perceived importance of using immunity certificates) as the dependent variable, and the independent variables described above. P-values smaller than 0.01 were used as threshold to indicate significance of the estimated coefficients. This analysis was performed in STATA[35]. Stepwise regression analysis was used in other studies exploring COVID-19 vaccination views[36,37], relationships between a COVID-19 risk index and COVID-19 mortality rates[38], anxiety and depression during COVID-19[27]. A graphical representation of the steps used in our statistical analysis is presented in Figure 3.

**Figure 3.**
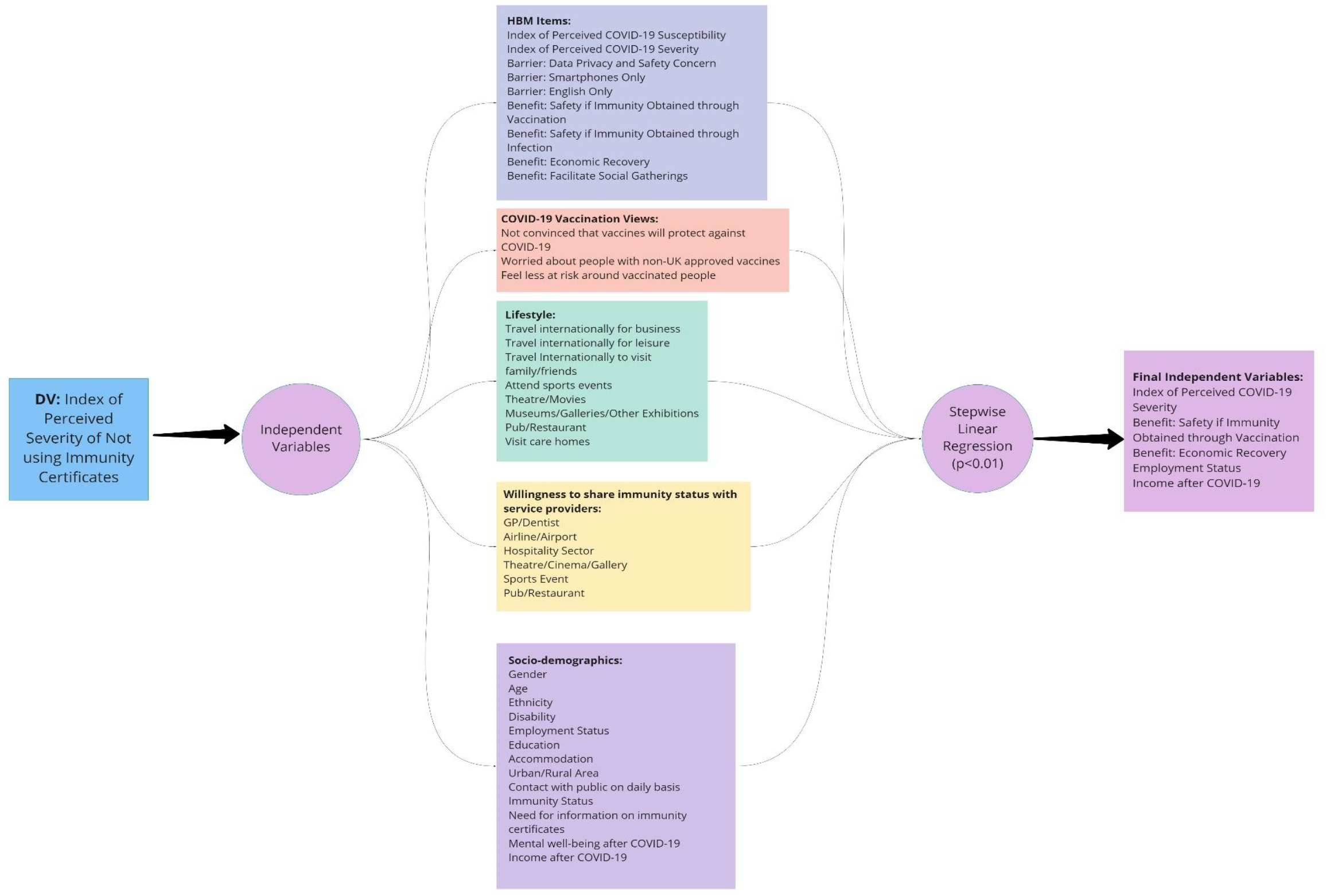
Illustration of the statistical analysis *(https://osf.io/jubv6)*

### 2.5. Power calculation

The sample size was chosen pragmatically based on several different approaches[39], obtaining a minimum sample size between 271 and 1,067 participants, depending on the assumptions.

## 3. Results

Table 3 presents our statistical model after conducting the multiple stepwise linear regression analysis with p<0.01. Respondents who perceived themselves as being more at risk of experiencing negative health consequences if they contracted the virus (*Perceived COVID-19 Severity*) were more likely to value positively the importance of immunity certificates (*Certificate Severity*) demonstrated with an increase of 0.2506 units (Table 3). Figure 4 illustrates the relationship between perceived importance of using immunity certificates and *Perceived COVID-19 Severity*.

**Table 3.** Stepwise Linear Regression Results of Certificate Severity and Perceived COVID-19 Severity, Benefit: Safe if Immunity Obtained through Vaccination, Benefit: Economic Recovery, Employed/Self-Employed, Income after COVID-19

| | $\beta$ | Std. Error | t-statistic | P>\|t\| | [95% conf. interval] | |
| --- | --- | --- | --- | --- | --- | --- |
| Perceived COVID-19 Severity | 0.2506*** | 0.0505 | 4.9600 | 0.0000 | 0.1513 | 0.3498 |
| Benefit: Safe if Immunity Obtained through Vaccination | 0.1594*** | 0.0325 | 4.9000 | 0.0000 | 0.0955 | 0.2233 |
| Benefit: Economic Recovery | 0.1585*** | 0.0344 | 4.6100 | 0.0000 | 0.0909 | 0.2261 |
| Employed/Self-employed | -0.2343*** | 0.0715 | -3.2800 | 0.0010 | -0.3747 | -0.0939 |
| Income after COVID-19 | -0.1280*** | 0.0408 | -3.1400 | 0.0020 | -0.2082 | -0.0478 |
| (Constant) | 1.6911*** | 0.2292 | 7.3800 | 0.0000 | 1.2408 | 2.1414 |

*Note: The adjusted R-squared of this regression is 22.76%. Employed/Self-Employed is a dummy variable that equals 1 if the respondent was either employed or self-employed at the time of the survey, and 0 if they are either retired or unemployed. \*\*\* $p < 0.01$ , \*\* $p < 0.05$ , \* $p < 0.1$ .*

**Figure 4.**
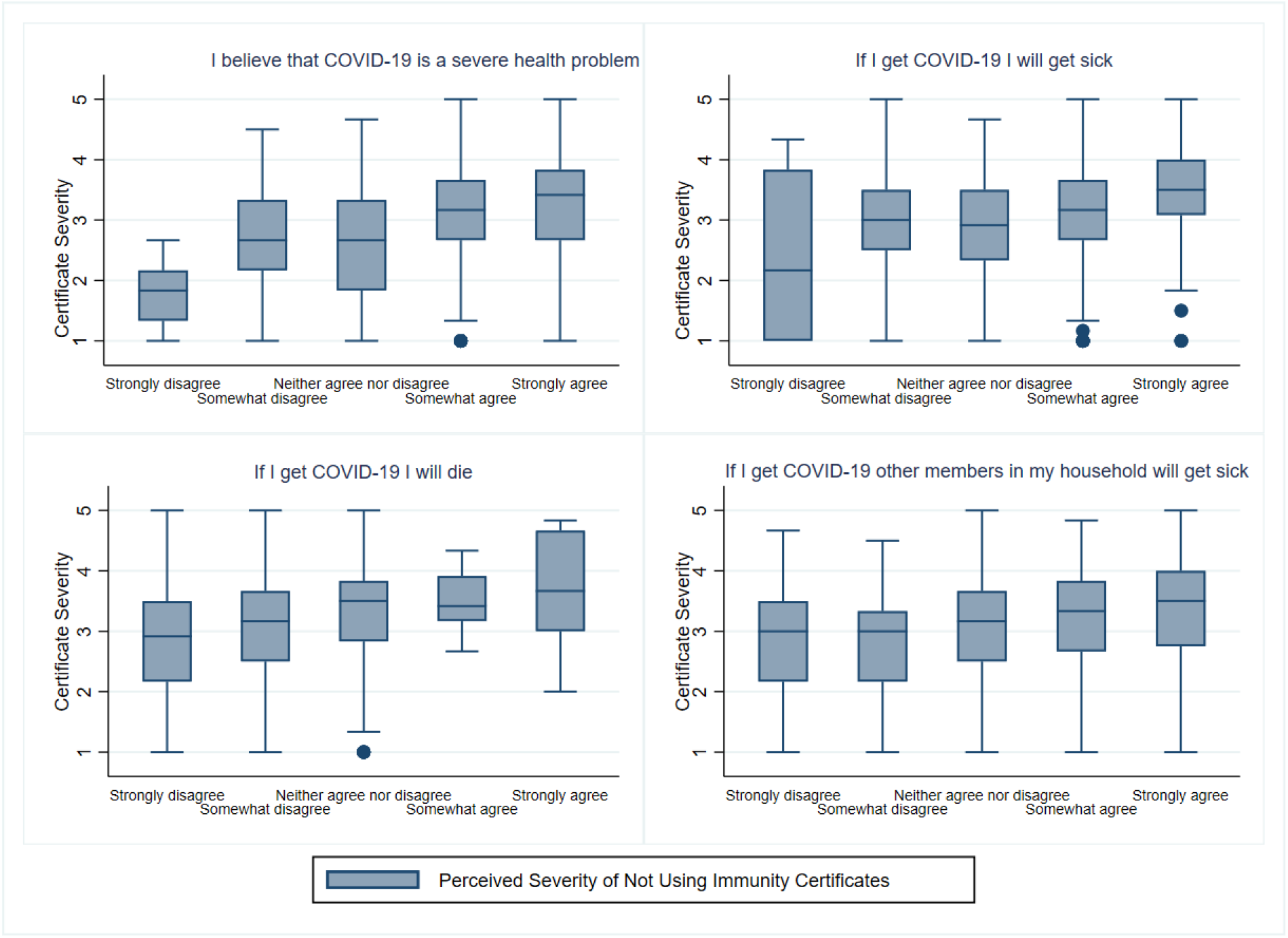
Perceived Importance of Using Immunity Certificates (*Certificate Severity)* by *Perceived COVID-19 Severity*.

Similarly, those who felt safer if vaccinated and believed in the prospect of future economic recovery were more likely to perceive as more important the use of immunity certificates, demonstrated with an increase of 0.1594 and 0.1585 units in *Certificate Severity* respectively (Table 3). Also, the results showed that those who were employed/self-employed or had experienced an increase in their net income after the COVID-19 outbreak were more likely to perceive as less important the use of immunity certificates. Specifically, compared to respondents who were retired or unemployed, those who were in employment (employed/self-employed) displayed a lower perceived importance of using immunity certificates (*Certificate Severity*) by 0.2343 units. The same negative effect was observed for people who reported higher levels of net income after the COVID-19 outbreak with a decrease of 0.1280 units in *Certificate Severity*. The relationship between perceived importance of using immunity certificates, employment status and net income after COVID-19 is presented in Figure 5.

**Figure 5.**
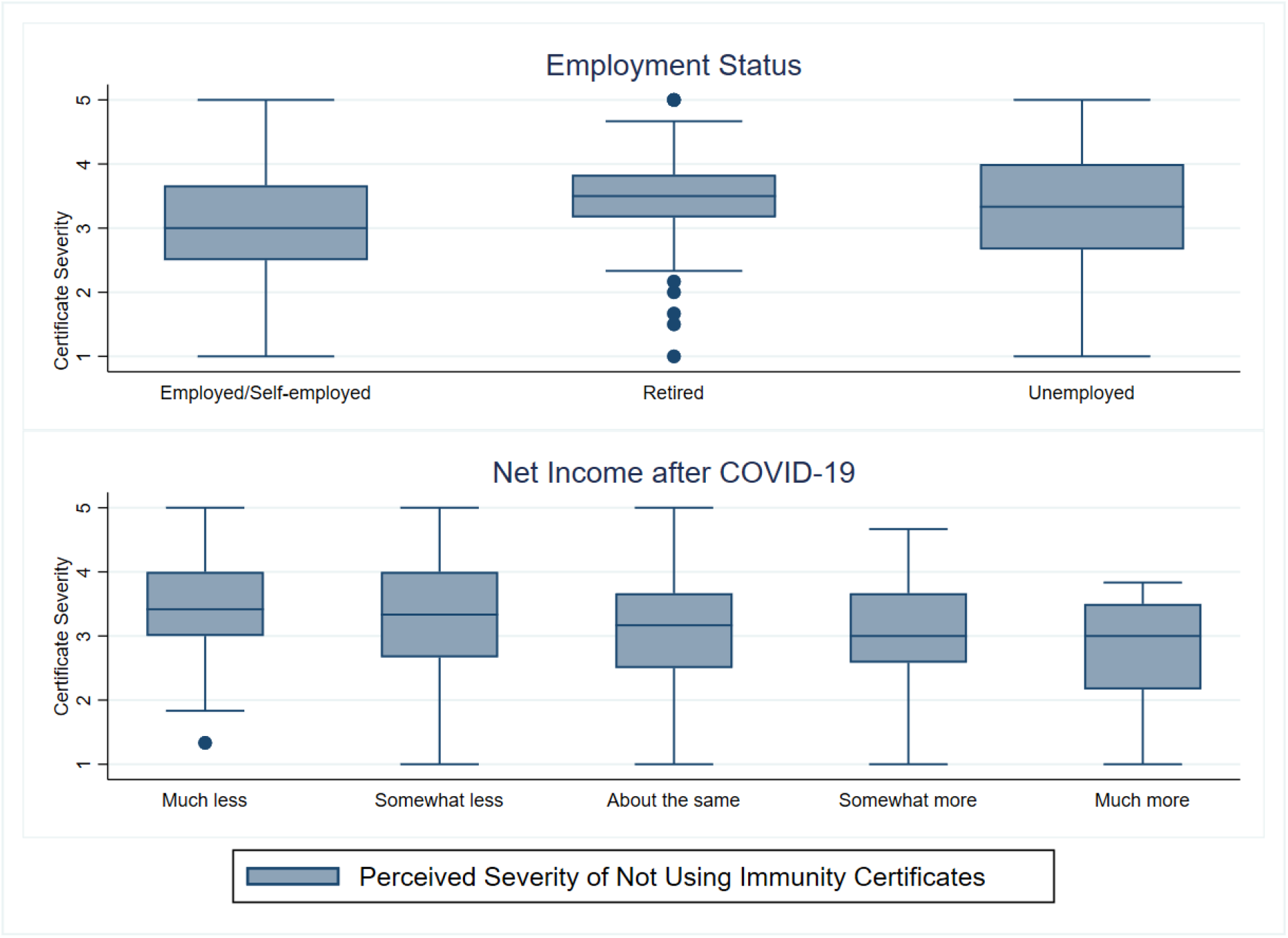
Perceived Importance of Using Immunity Certificates (*Certificate Severity)* by Employment Status and Net Income After COVID-19.

Finally, the remaining independent variables used in the statistical analysis including *Perceived COVID-19 Susceptibility*, lifestyle, age, gender and ethnicity (among others) did not have a statistically significant effect on perceived importance of using immunity certificates.

## 4. Discussion

The findings of our research suggest that people who are more vulnerable (not working and believing that contracting COVID-19 would have a severe impact on their health) are more responsive to the use of immunity certificates and therefore the importance of using them in daily life is perceived as higher. Also, respondents perceived the importance of immunity certificates as higher if immunity was acquired after a full course of vaccination compared to past infection. These findings partially confirm the results of previous studies where the authors investigated the role of personal health beliefs on vaccination[1–3]. Also, as opposed to previous research on attitudes towards vaccination we did not find an effect of age, gender and ethnic background when it comes to the perceived importance of immunity certificates[1,2,23]. However, we did observe significant effect of employment status and loss of income suggesting the importance of socio-economic factors compared to demographics in this context.

### Limitations

One of the limitations of our study is that participants were recruited from the online survey platform Prolific. Since Prolific surveys are completed digitally (mobile, PC, tablet etc.) our sample was comprised of people who had the means and capacity to use digital technologies.

Another limitation of our study is the relatively low explanatory power of our model with and adjusted R-squared of 22.76%, suggesting that the independent variables chosen by our stepwise linear regression model only explain 22.76% of the observed variation in the index *Certificate Severity*. Considering that research on immunity certificates is still in its early stages, we do not yet have a large body of literature to draw from in order to identify more predictors of *Certificate Severity*. More research is needed to explore what the factors that we do not capture could be.

## 5. Conclusions

Understanding the role of individual factors on the perceived importance of immunity certificates is necessary to make evidence-based decisions when considering their design and implementation. Such decisions should aim to protect vulnerable members of our society and those in need.

## Data Availability

The data is available in a public, open access repository. All materials are freely available on OSF (https://osf.io/jubv6/)

## Footnotes

### Contributors

The questionnaire survey was conceptualised by CN, IS and PB, with the input of TG, CLA, and OC. CEN and PB completed the data collection. CN and IS conducted the statistical analysis. All authors contributed and approved the final manuscript.

### Ethics statements

Ethics approval was obtained from the College of Engineering, Design and Physical Sciences Research Ethics Committee at Brunel University London (Ref. 31705-A-Jul/2021-33586-1) on the 29^th^ of July 2021.Informed consent was obtained from all respondents prior to the beginning of the survey. Respondents were allowed to withdraw from the survey at any time.

### Data availability statement

The data is available in a public, open access repository. All materials are freely available on OSF *(https://osf.io/jubv6/)*

